# Evaluation of paraspinal muscle quality using Hounsfield unit in simple elliptical regions of interest: correlation with magnetic resonance imaging-based intramuscular fat infiltration in spine surgery patients

**DOI:** 10.64898/2026.08.31.26361574

**Authors:** Naoki Segi, Yuya Okada, Yosuke Takeichi, Sadayuki Ito, Jun Ouchida, Yasuhiro Nagatani, Yujiro Kagami, Hiroto Tachi, Kazuma Ohshima, Keisuke Ogura, Shiro Imagama, Hiroaki Nakashima

## Abstract

**Study design:** Retrospective cohort study.

**Objectives:** To correlate Hounsfield unit (HU) values, using elliptical regions of interest (ROI), that can be easily defined in routine clinical practice with magnetic resonance imaging (MRI) T2-hyperintense area fraction, as a surrogate for paraspinal muscle fat infiltration and to establish specific HU screening thresholds that may be applied with standard picture archiving and communication system (PACS).

**Methods:** We included 136 patients (71 men; 61.0 ± 15.4 years) who underwent preoperative computed tomography (CT) and MRI within an 8-week period. Elliptical ROI HU values were measured at L2/3 and L4/5 for erector spinae, multifidus, and psoas major. MRI T2-hyperintense area fraction (Otsu thresholding) served as the fat infiltration reference. Linear mixed-effects (LME) models were used to assess the HU–T2 association and level-specific receiver operating characteristic (ROC) analyses (lower HU value side; n=136 per muscle–level) to identify thresholds for ≥30% and ≥50% infiltration criteria.

**Results:** Intraclass coefficients = 0.709 (HU) and 0.857 (T2 fraction); Goutallier weighted kappa = 0.579. In the overall LME, β was −0.880 HU per 1% T2-fraction increase (95% confidence interval −0.935 to −0.825; marginal R² =0.502); the association was steeper in multifidus (β = −1.020) than in erector spinae (β = −0.753). Psoas major (*R* = −0.226) was excluded from ROC analyses. Difference between L2/3 and L4/5 HU cutoffs was ∼20 HU. The ≥50% criterion revealed higher discrimination.

**Conclusions:** Elliptical ROI-based HU measurements may reliably screen paraspinal muscle fat infiltration in erector spinae and multifidus using standard PACS. Specific thresholds may allow practical preoperative evaluation without additional costs or radiation.

## Introduction

Degenerative spinal disease is one of the leading causes of disability worldwide, and the number of patients undergoing lumbar spine surgery is continuously increasing.^1^ In addition to advances in surgical technique and implant technology, preoperative patient characterization plays an essential part in risk stratification and outcome prediction. Among the factors that have gained attention in this context, paraspinal muscle quality, particularly intramuscular fat infiltration, has emerged as an important determinant of surgical outcomes, including functional recovery, postoperative pain, fusion success, adjacent segment degeneration, proximal junctional kyphosis, and pedicle screw loosening.^2,3^ Furthermore, muscle degeneration associated with fat infiltration has been closely linked to the broader concepts of sarcopenia and frailty, which can independently predict perioperative complications, prolonged hospitalization, and reduced quality-of-life improvement after spine surgery.^4,5^ Therefore, accurate preoperative assessment of paraspinal muscle quality is of substantial clinical value.

Currently, two principal approaches are used to evaluate intramuscular fat infiltration. The first approach is the Goutallier classification, originally described for computed tomography (CT)-based assessment of rotator cuff musculature^6^ and subsequently adapted for MRI-based evaluation of lumbar paraspinal muscles. Despite its widespread adoption, the Goutallier grading system carries recognized limitations including moderate inter-observer agreement (typically weighted κ ≈ 0.5–0.7)^7,8^ and Grade 2 spanning a broad range of fat infiltration (25%–50%), which may potentially obscure clinically important differences. The second approach involves quantitative image-analysis software pipelines—such as thresholding algorithms applied to segmented freehand regions of interest (ROIs) on T1- or T2-weighted magnetic resonance imaging (MRI)—that yield continuous fat fraction estimates.^9,10^ Although such methods provide greater precision, they require dedicated image-processing software outside standard clinical picture archiving and communication system (PACS) environments, are time-consuming, and are rarely feasible outside dedicated research settings. Moreover, MRI signal intensity is inherently device-dependent and lacks absolute physical reference, precluding direct cross-institution comparison without normalization.^11^

Against this backdrop, Hounsfield unit (HU) measurement on routine CT offers a compelling alternative. HU values are physically defined by the linear attenuation coefficient of X-rays relative to water, making them intrinsically standardized across scanner vendors and imaging sites. CT is routinely performed as a part of preoperative planning for the majority of spinal surgery candidates, and HU values can be measured directly using standard PACS software without additional cost, processing time, or radiation. The value of HU-based assessment has been well established for opportunistic vertebral bone quality screening, where trabecular HU values at lumbar vertebrae have been validated as surrogates for dual-energy X-ray absorptiometry-measured bone mineral density.^12,13^ Accordingly, muscle HU values—which decrease with increasing intramuscular fat content due to the low attenuation of adipose tissue—have been used to characterize myosteatosis in sarcopenia and various systemic diseases.^14^ However, the relationship between muscle HU measured via simple, clinician-applicable elliptical ROIs and quantitative MRI-based intramuscular fat infiltration, specifically in lumbar paraspinal muscles, remains unelucidated.

Therefore, in this study, we aimed (1) to quantify the association between CT-based elliptical ROI HU values and MRI-based T2-hyperintense area fraction as a surrogate for intramuscular fat infiltration in the lumbar erector spinae, multifidus, and psoas major and (2) to determine level-specific HU thresholds that can screen clinically meaningful levels of fat infiltration (≥30% and ≥50%), providing practical cutoff values that can be applied with standard PACS software. By exploring a versatile method, for evaluating the quality of paraspinal muscles, that can be completed quickly within a PACS environment alone, we aimed to provide a foundation for a paraspinal muscle evaluation method that can be directly applied to daily clinical practice and inform future research.

## Materials and Methods

### Study Design and Patient Population

This retrospective study was conducted in accordance with the Declaration of Helsinki and approved by the institutional review board (approval number is listed separately). The requirement for individual informed consent was waived by the approving body because the study used only de-identified imaging data.

Patients who underwent elective spinal surgery with preoperative lumbar CT and MRI at our institution between January 2023 and December 2025 were screened for inclusion. Eligibility criteria comprised age ≥20 years and CT and MRI examinations of the lumbar spine performed within an 8-week interval. Exclusion criteria comprised prior lumbar spine surgery, skeletal dysplasia or metabolic bone diseases, and poor image quality of the CT/MRIs precluding reliable measurement.

### CT Image Acquisition and Measurement

CT examinations were performed using Aquilion Prime SP (Canon Medical Systems, Otawara, Japan) with standard lumbar spine protocols (tube voltage 120 kV; tube current modulated automatically, approximately 200–300 mA). All CT data were analyzed using SYNAPSE SAI Viewer FS-V686 (FUJIFILM Corporation, Tokyo, Japan). Axial images at the mid-level of the L2/3 and L4/5 intervertebral disc spaces were identified. At each level, bilateral erector spinae, multifidus, and psoas major were measured using the built-in elliptical ROI tool. The ellipse was positioned to maximally inscribe the visible muscle cross-section, while excluding adjacent muscles, fascial planes, and any sharply demarcated focal signal abnormalities at its periphery. The ellipse axes were not rotated from the default image x/y orientation. The software-reported mean HU values were recorded (Figure 1). All measurements were performed by a single observer blinded to MRI findings, and the measurements were re-performed for a random subset of 30 patients by a second blinded observer to assess inter-observer reliability. Further, the HU values were measured using freehand ROIs drawn along the apparent muscle boundaries using an optical mouse (MS116p; Dell Technologies Inc., Round Rock, TX, USA).

**Figure 1.**
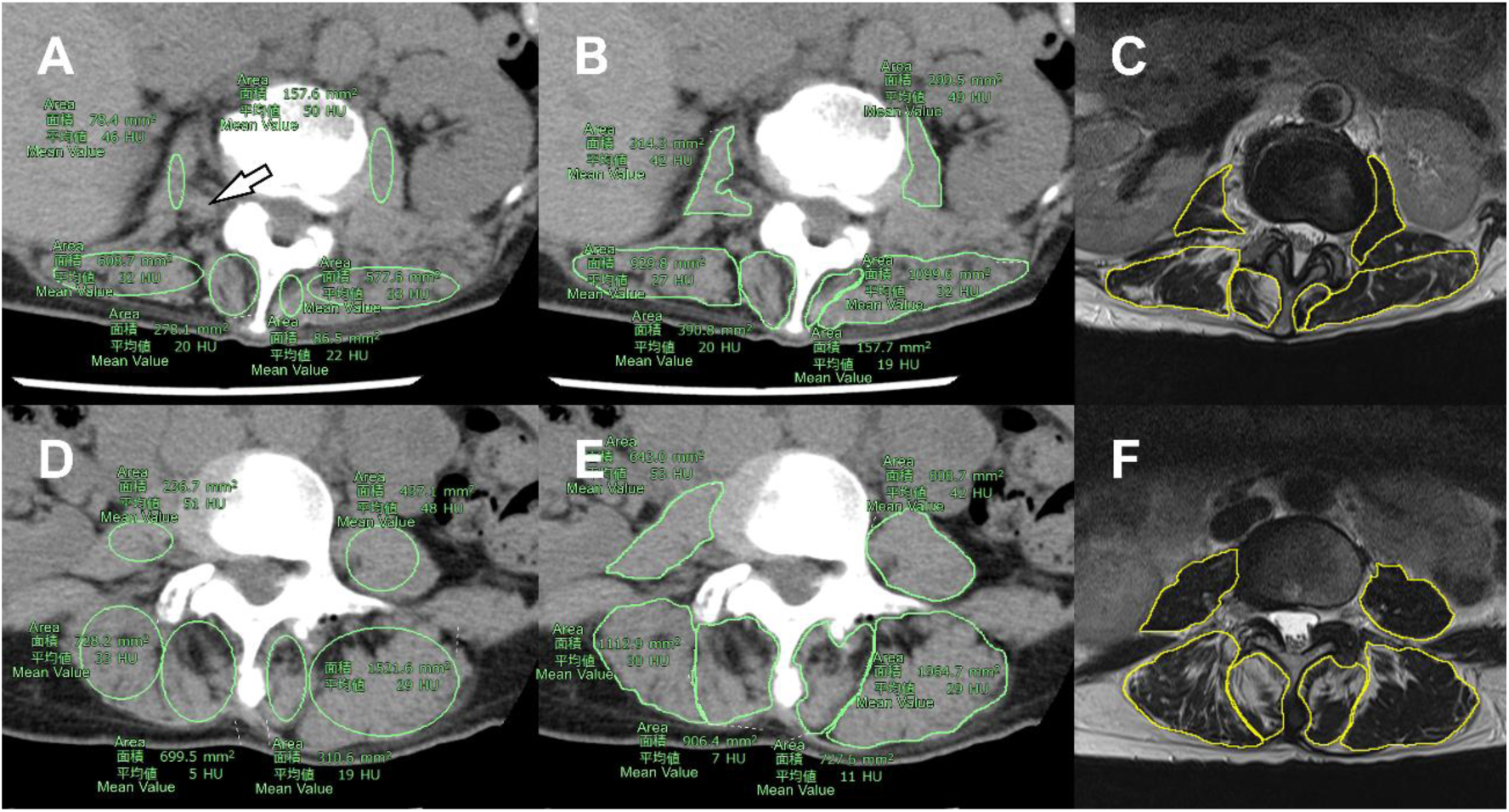
Application of measuring HU values and T2-hyperintense area fraction of paraspinal muscles. A. Placement of the elliptical region of interest (ROI) on axial computed tomography (CT) image at the L2/3 level. The ellipse was positioned to maximally inscribe the visible muscle cross-section, while excluding adjacent muscles, fascial planes, and any sharply demarcated focal signal abnormalities at its periphery. The ellipse axes were not rotated from the default image x/y orientation. Note that in the right psoas major, the low-density area adjacent to the vertebral body was considered to be perineural fat and was therefore excluded from the ROI (arrow). B. Placement of manually traced freehand ROIs on the same image. C. Placement of manually traced freehand ROIs for the T2-hyperintense area fraction measurements on the T2-weighted transverse magnetic resonance image corresponding to that in panel A. D–F show similar examples at the L4/5 level.

### MRI Image Acquisition and Measurement

MRI examinations were performed using one of the following clinical scanners: Magnetom Aera, Magnetom Avanto.fit (1.5 T; Siemens Healthineers, Erlangen, Germany), Magnetom Skyra.fit (3 T; Siemens Healthineers), Vantage Centurian (3 T; Canon Medical Systems Corporation, Otawara, Japan), or Ingenia Prodiva CX (1.5 T; Philips Healthcare, Amsterdam, Netherlands). Axial T2-weighted fast spin-echo sequences were acquired with repetition time 3,000–7,000 ms and echo time 80–120 ms (specific parameters varied by scanner and clinical protocol). Multiple scanners were used to mimic routine clinical practice. Notably, image quality was confirmed to be adequate for muscle boundary delineation in all included cases. DICOM data were exported in de-identified form and analyzed offline using Fiji 1.54p (ImageJ2; National Institutes of Health, Bethesda, MD, USA).^15^ Axial T2-weighted images at the mid-level of the L2/3 and L4/5 disc spaces were identified to correspond to the CT measurements to the extent possible.

For each muscle at each level, a freehand ROI was manually traced along the apparent muscle boundary using a pen tablet (Intuos Pro 4; Wacom Co., Ltd., Kazo, Japan). A two-class Otsu threshold^16^ was applied to the grayscale histogram to separate high-signal from low-signal components, and the proportion of pixels exceeding the threshold was recorded as the T2-hyperintense area fraction (%). This fraction served as the primary MRI-based surrogate for intramuscular fat infiltration; however, notably, T2 hyperintensity may also reflect edema, denervation, and other non-lipid changes.^9^ In addition, each muscle was graded according to a modified Goutallier classification (Grade 0: no fatty signal; Grade 1: some fatty streaks; Grade 2: <50% fatty signal; Grade 3: 50% fatty signal; Grade 4: >50% fatty signal).^6,7^ MRI measurements were performed by the same observers who performed the CT measurements; however, the CT results were inaccessible during the MRI assessment.

### Statistical Analysis

Statistical analyses were performed using R (version 4.6.0 (2026-04-24 ucrt); R Core Team, Vienna, Austria) with packages tidyverse (version 2.0.0), gtsummary (version 2.5.1), irr (version 0.84), lme4 (version 2.0.1), sjPlot (version 2.9.0), pROC (version 1.19.0.1). Inter-observer reliability for HU and T2-hyperintense fraction was quantified using two-way mixed-effects intraclass correlation coefficients (ICC; absolute agreement, single measure). Inter-observer reliability for Goutallier grading was quantified using linearly weighted Cohen’s kappa.

Of the 1,632 observations (136 patients × 3 muscles × 2 levels × 2 sides), after excluding three observations because they were deemed unmeasurable and resulted in missing values, 1,629 observations were analyzed. To assess the association between the HU and T2-hyperintense fraction, while accounting for the hierarchical data structure (multiple muscles and levels measured per patient), linear mixed-effects models (LME) were fitted, with patient identity as a random intercept. Left and right sides were retained as separate observations to preserve clinically relevant lateral asymmetry, with side included as a fixed covariate. Further, muscle type, spinal level, and T2-hyperintense fraction were included as fixed effects in the overall model. Muscle-specific models for erector spinae and multifidus were fitted separately. The marginal coefficient of determination (R²_m, representing variance explained by fixed effects alone) and conditional R² (R²_c, including the random effect) were computed.

For level-specific receiver operating characteristic (ROC) analyses, the side with the lower HU value per patient per muscle per level was selected, yielding 136 independent observations per muscle-level combination. Given that previous studies have indicated that a value >30% in the T2-hyperintense fraction of the paraspinal muscles is associated with poor postoperative outcomes,^17^ and that the most severe grade in the Goutallier classification, grade 4, is defined as >50%, ROC curves were constructed with HU as the predictor for binary classification of T2-hyperintense fraction ≥30% and ≥50%. The Youden index was used to identify optimal HU cutoffs. Area under curve (AUC) 95% confidence intervals (CIs) were computed by the DeLong method. Statistical significance was set at *P* < 0.05. However, because the psoas major muscle revealed only a weak overall correlation and the prevalence of fat infiltration was extremely low, class imbalance made it impossible to perform a reliable ROC analysis. Therefore, it was excluded from the threshold analysis and described separately.

## Results

Overall, 136 patients were enrolled (71 men, 65 women; mean age 61.0 ± 15.4 years; range 21–94 years) in this study. There was no significant difference in mean age between men and women. Men had a significantly higher body mass index (25.0 ± 4.6 vs. 23.7 ± 5.6; *P* = 0.049). The most common diagnosis was schwannoma (23.5%), followed by lumbar spinal stenosis (20.6%) and spinal deformity (14.0%); 13.2% of patients had non-lumbar spinal diagnoses, such as stenosis at the lower thoracic levels (Table 1). The interval between CT and MRI examinations was 22.0 ± 16.4 days (range 0–52 days).

**Table 1.** Baseline demographic and clinical characteristics of the study population.

|  | **Overall**<br>N = 136 | **Men**<br>n = 71 | **Women**<br>n = 65 | **P value** |
| --- | --- | --- | --- | --- |
| **Age** |  |  |  | 0.17 |
| Mean ± SD | 61.0 ± 15.4 | 60.2 ± 14.5 | 61.8 ± 16.4 | |
| Min, Max | 21, 94 | 23, 94 | 21, 86 |  |
| **Height** | 160.0 ± 10.2 | 166.3 ± 7.0 | 153.1 ± 8.6 | **<0.001** |
| **Weight** | 62.8 ± 16.0 | 69.6 ± 16.0 | 55.4 ± 12.4 | **<0.001** |
| **BMI** | 24.4 ± 5.2 | 25.0 ± 4.6 | 23.7 ± 5.6 | **0.049** |
| **Diagnosis** |  |  |  | **0.016<sup>†</sup>** |
| Schwannoma | 32 (23.5%) | 20 (28.2%) | 12 (18.5%) |  |
| LCS | 28 (20.6%) | 20 (28.2%) | 8 (12.3%) |  |
| ASD | 19 (14.0%) | 6 (8.5%) | 13 (20.0%) |  |
| No disease | 18 (13.2%) | 11 (15.5%) | 7 (10.8%) |  |
| Listhesis | 12 (8.8%) | 5 (7.0%) | 7 (10.8%) |  |
| LDH | 11 (8.1%) | 5 (7.0%) | 6 (9.2%) |  |
| Fracture | 6 (4.4%) | 1 (1.4%) | 5 (7.7%) |  |
| AIS | 5 (3.7%) | 0 (0.0%) | 5 (7.7%) |  |
| Other | 5 (3.7%) | 3 (4.2%) | 2 (3.1%) |  |

<sup>†</sup> Pearson's Chi-squared test with simulated p-value (based on 2000 replicates).
LCS, lumbar canal stenosis; ASD, adult spinal deformity; LDH, lumbar disc herniation; AIS, adolescent idiopathic scoliosis.

The elliptical ROI-based HU values were lowest in the multifidus muscles at both the L2/3 and L4/5 levels (39.8 ± 25.3 and 25.6 ± 33.7, respectively) and highest in the psoas major muscles (48.2 ± 10.7 and 49.8 ± 9.0, respectively). While the erector spinae and multifidus muscles could exhibit negative HU values, the psoas major muscle could not. The proportion of muscle with T2-hyperintense fraction ≥30% was 31.3% (L2/3) and 41.2% (L4/5) for multifidus, compared with 3.3% (L2/3) and 1.5% (L4/5) for psoas major. The proportion of muscle with T2-hyperintense fraction ≥50% was 12.1% (L2/3) and 17.3% (L4/5) for multifidus, compared with 1.5% (L2/3) and 0% (L4/5) for psoas major. At both L2/3 and L4/5 levels, Grade 1 (some fatty streaks) was the most common Goutallier category for erector spinae and multifidus, whereas Grade 0 (no fatty infiltration) was the most common category for psoas major (Table 2).

**Table 2.** Paraspinal muscle quality characteristics at L2/3 and L4/5 levels by muscle type.

|  | L2/3 |  |  | L4/5 |  |  |
| --- | --- | --- | --- | --- | --- | --- |
|  | Erector<br>spinae<br>n = 272 | Multifidus<br>n = 272 | Psoas<br>major<br>n = 272 | Erector<br>spinae<br>n = 272 | Multifidus<br>n = 272 | Psoas<br>major<br>n = 272 |
| **Elliptical ROI HU value** |  |  |  |  |  |  |
| Mean ± SD | 42.5 ± 15.2 | 39.8 ± 25.3 | 48.2 ± 10.7 | 33.1 ± 22.2 | 25.6 ± 33.7 | 49.8 ± 9.0 |
| Min, Max | -49, 66 | -66, 71 | 7, 79 | -58, 73 | -71, 71 | 11, 88 |
| Unable | 0 | 0 | 2 | 0 | 0 | 1 |
| **MRI T2-hyperintense fraction, %** |  |  |  |  |  |  |
| | 13.7 ± 13.7 | 23.5 ± 19.9 | 6.2 ± 9.8 | 18.9 ± 16.9 | 29.4 ± 20.8 | 4.6 ± 6.6 |
| Unable | 0 | 0 | 0 | 0 | 0 | 1 |
| 30%< | 24 (8.8%) | 85 (31.3%) | 9 (3.3%) | 56 (20.6%) | 112 (41.2%) | 4 (1.5%) |
| 50%< | 8 (2.9%) | 33 (12.1%) | 4 (1.5%) | 17 (6.3%) | 47 (17.3%) | 0 (0.0%) |
| **Goutallier classification** |  |  |  |  |  |  |
| 0 | 108<br>(39.7%) | 103<br>(37.9%) | 224<br>(82.7%) | 72 (26.5%) | 38 (14.0%) | 214<br>(79.0%) |
| 1 | 116 (42.6%) | 94 (34.6%) | 28 (10.3%) | 100 (36.8%) | 80 (29.4%) | 43 (15.9%) |
| 2 | 36 (13.2%) | 39 (14.3%) | 16 (5.9%) | 64 (23.5%) | 67 (24.6%) | 13 (4.8%) |
| 3 | 8 (2.9%) | 13 (4.8%) | 2 (0.7%) | 27 (9.9%) | 38 (14.0%) | 1 (0.4%) |
| 4 | 4 (1.5%) | 23 (8.5%) | 1 (0.4%) | 9 (3.3%) | 49 (18.0%) | 0 (0.0%) |
| Unable | 0 | 0 | 1 | 0 | 0 | 1 |

ROI, region of interest; HU, Hounsfield unit; MRI, magnetic resonance imaging.

Inter-observer ICC was 0.709 for elliptical HU values and 0.857 for T2-hyperintense fraction. Inter-observer reliability for Goutallier grading (weighted kappa) was 0.579 (Table 3). The correlation coefficient between HU values derived from freehand ROIs and those derived from elliptical ROIs was 0.776 (ICC, 0.733) (Supplemental Figures 1 and 2). The time required to measure the HU value at a single location was 24.3 ± 13.2 s when using a freehand ROI and 4.0 ± 1.4 s when using an elliptical ROI.

### Unadjusted Association between HU and T2-Hyperintense Fraction

Treating all measurements as independent observations, HU values negatively correlated with T2-hyperintense fraction across all muscles, levels, and sides (Pearson *R*, −0.729; 95% CI, −0.747 to −0.710; *P* < 0.001; Figure 2). The correlation was consistent across sexes (men: *R*, −0.687; women: *R,* −0.717; both *P* < 0.001) and age groups (<65 years: *R*, −0.675; 65–74 years: *R*, −0.662; ≥75 years: *R*, −0.726; all *P* < 0.001). By muscle type, correlations were strong for erector spinae (*R,* −0.713) and multifidus (*R*, −0.745), but distinctly weak for psoas major (*R*, −0.226; all *P* < 0.001).

**Figure 2.**
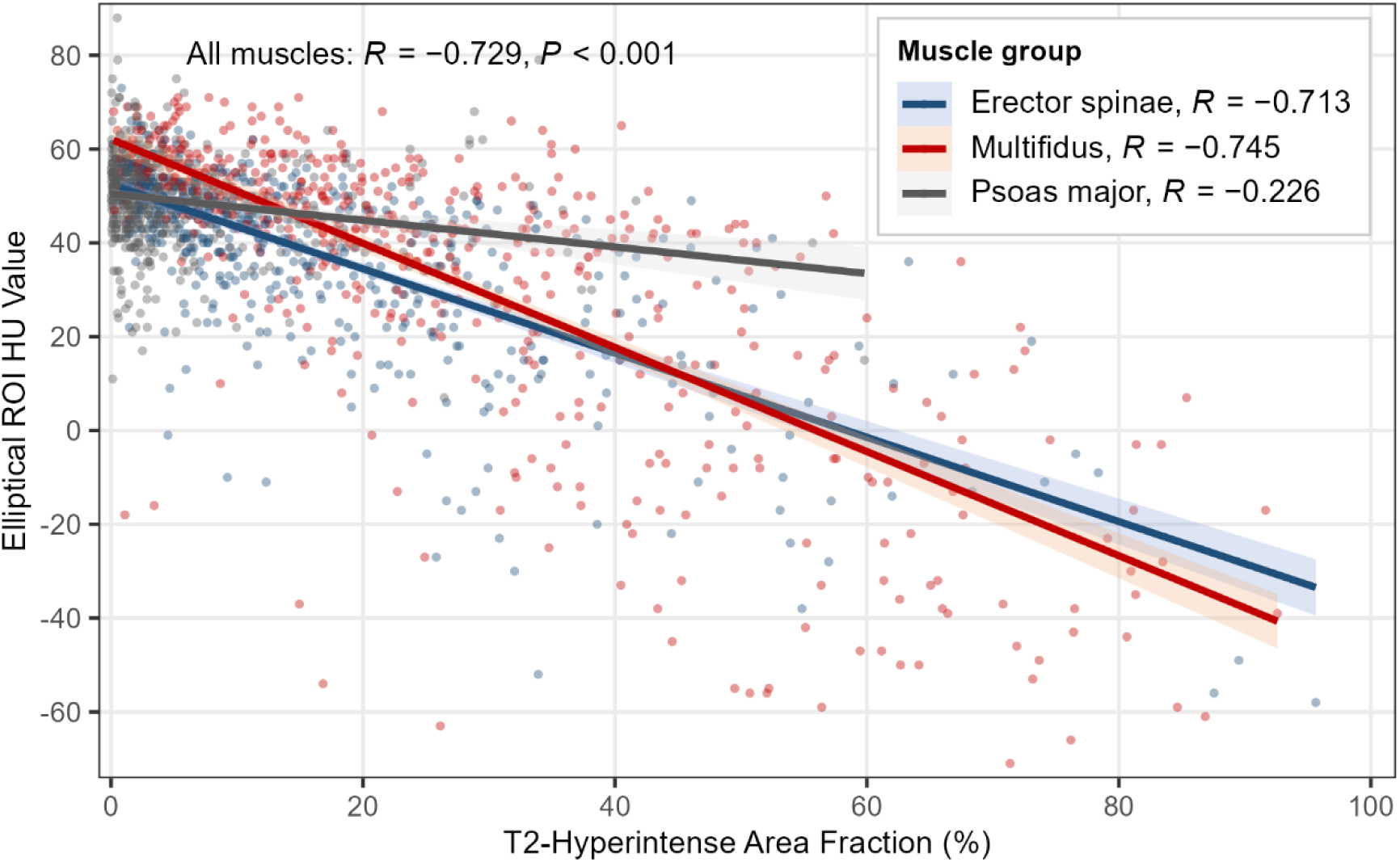
Unadjusted association between elliptical ROI HU value and MRI T2-hyperintense area fraction across all paraspinal muscles. Solid lines indicate ordinary least-squares regression fits for each muscle type, with shaded bands representing 95% confidence intervals. Pearson correlation coefficients are annotated for each muscle group: erector spinae *R* = −0.713, multifidus *R* = −0.745, psoas major *R* = −0.226 (all *P* < 0.001); the overall correlation across all muscle types was *R* = −0.729 (*P* < 0.001). Note that these coefficients treat all observations independently; adjusted estimates from linear mixed-effects models are reported in Table 4 and the main text. ROI, region of interest; HU, Hounsfield unit; MRI, magnetic resonance imaging.

**Table 3.** Inter-observer reliability of CT, MRI, and qualitative muscle assessments.

|  | **Kappa** | **ICC** | **95%CI** | ***P* value** |
| --- | --- | --- | --- | --- |
| Goutallier classification | 0.579 |  |  | <0.001 |
| Elliptical ROI HU value |  | 0.709 | 0.603–0.782 | <0.001 |
| Elliptical vs freehand ROI HU value |  | 0.733 | 0.565–0.825 | <0.001 |
| MRI T2-hyperintense fraction |  | 0.857 | 0.756–0.909 | <0.001 |

CT, computed tomography; MRI, magnetic resonance imaging; ICC, intraclass correlation coefficient; CI, confidence interval; ROI, region of interest; HU, Hounsfield unit.

**Table 4.** Linear mixed-effects model results for the association between elliptical ROI HU values and MRI T2-hyperintense fraction.

| Parameter | Overall model | Erector spinae | Multifidus |
| --- | --- | --- | --- |
| **Fixed effects, $\beta$ [95%CI]** |  |  |  |
| Intercept (HU)<sup>†</sup> | 54.774<br>[52.650 to 56.899] | 53.104<br>[50.650 to 55.558] | 64.592<br>[60.729 to 68.455] |
| T2-high fraction (per 1%) | −0.880*<br>[−0.935 to −0.825] | −0.753*<br>[−0.843 to −0.663] | −1.020*<br>[−1.121 to −0.920] |
| Level, L4/5 vs L2/3 (HU) | −4.498*<br>[−5.824 to −3.173] | −5.622*<br>[−7.416 to −3.828] | −8.203*<br>[−10.930 to −5.475] |
| Side, right vs left (HU) | −0.769<br>[−2.083 to +0.546] | −0.459<br>[−2.194 to +1.275] | −1.528<br>[−4.197 to +1.141] |
| Muscle, multifidus vs erector<sup>‡</sup> | +3.877*<br>[+2.175 to +5.579] | — | — |
| Muscle, psoas vs erector<sup>‡</sup> | +1.483<br>[−0.238 to +3.204] | — | — |
| **Random effects** |  |  |  |
| Patient-level variance | 61.62 | 82.15 | 155.99 |
| Residual variance | 182.66 | 105.86 | 250.04 |
| Patient-level ICC | 0.25 | 0.38 | 0.36 |
| **Model fit** |  |  |  |
| Marginal R<sup>2</sup> | 0.502 | 0.454 | 0.543 |
| Conditional R<sup>2</sup> | 0.628 | 0.693 | 0.719 |
| Observations | 1,629 | 544 | 544 |
| Patients | 136 | 136 | 136 |

<sup>†</sup> Intercept represents estimated HU at intramuscular T2-high area = 0%, spinal level L2/3, left side, and erector spinae in the overall model.
<sup>‡</sup> Muscle type fixed effects appear in the overall model only; muscle-specific models include only the respective muscle.
ROI, region of interest; HU, Hounsfield unit; MRI, magnetic resonance image; CI, confidence interval. ICC, intraclass correlation coefficient (patient-level random effect variance / total variance). Marginal R<sup>2</sup> reflects the variance explained by fixed effects alone; conditional R<sup>2</sup> includes the patient random intercept.
\* P < 0.001; no asterisk means not significant.

### Adjusted Analysis (Linear Mixed-Effects Models)

In the overall LME accounting for the hierarchical data structure, T2-hyperintense fraction was a significant negative predictor of HU (β, −0.880 HU per 1% increase; 95% CI, −0.935 to −0.825; *P* < 0.001; marginal R², 0.502; conditional R², 0.628). The patient-level intraclass correlation was 0.25, indicating that 25% of total variance resided at the patient level; most variance was attributable to within-patient factors (muscle type, spinal level) that were explicitly modeled as fixed effects. After adjusting for muscle type and side, L4/5 measurements were systematically lower than L2/3 measurements by 4.5 HU (*P* < 0.001), independently of fat infiltration. No significant systematic difference was found between left- and right-side measurements (β = −0.769 HU; *P* = 0.252) (Table 4).

In muscle-specific models for erector spinae and multifidus, T2-hyperintense fraction remained a significant predictor (Table 4). The association was steeper in multifidus (β, −1.020; 95% CI, −1.121 to −0.920; R²_m, 0.543; R²_c, 0.719) than in erector spinae (β, −0.753; 95% CI, −0.843 to −0.663; R²_m, 0.454; R²_c, 0.693). In both models, L4/5 showed substantially lower HU than did L2/3, independently of T2-hyperintense fraction (erector spinae, −5.6 HU; multifidus, −8.2 HU; both *P* < 0.001); no significant left-right difference was observed.

Because class imbalance due to the near-absence of fat infiltration ≥30% in psoas major precluded reliable ROC analysis, psoas major was excluded from threshold analyses.

### Level-Specific HU Thresholds

Level-specific ROC analyses using the side with the lower HU value per patient (n = 136 per muscle-level combination) identified HU thresholds that differed between spinal levels (Table 5; Figures 3 and 4). For the ≥30% fat infiltration criterion, optimal HU cutoffs differed by approximately 20 HU between the L2/3 and L4/5 levels; erector spinae 35.5 (L2/3) vs. 17.0 (L4/5) and multifidus 28.5 (L2/3) vs. 9.5 (L4/5). AUC values ranged 0.865–0.903 for this criterion (Figure 3). For the ≥50% criterion, level differences were attenuated (erector spinae, 26.5 [L2/3] vs. 12.5 [L4/5] HU; multifidus, 24.5 [L2/3] vs. 9.5 [L4/5] HU), indicating higher discriminatory power (Table 5).

**Figure 3.**
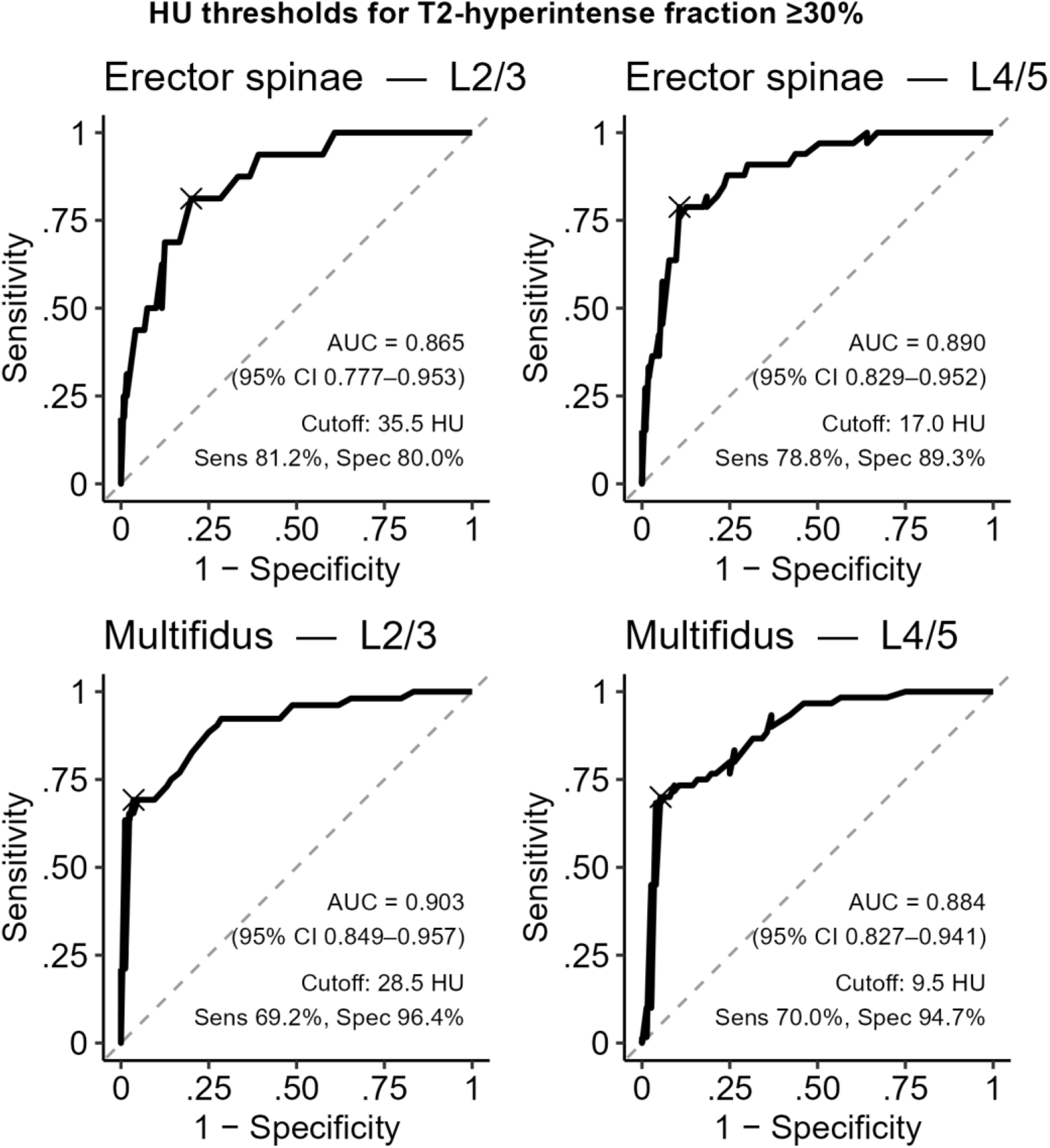
Level-specific receiver operating characteristic curves for detecting intramuscular T2-hyperintense area fraction ≥30%. Each analysis used the lower HU value side per patient, yielding n = 136 independent observations per panel. The “×” marks in each panel marks the Youden-optimal cutoff point. AUC with 95% confidence intervals (DeLong method), optimal HU cutoff, sensitivity, and specificity are annotated within each panel. HU, Hounsfield unit; AUC, area under the curve.

**Figure 4.**
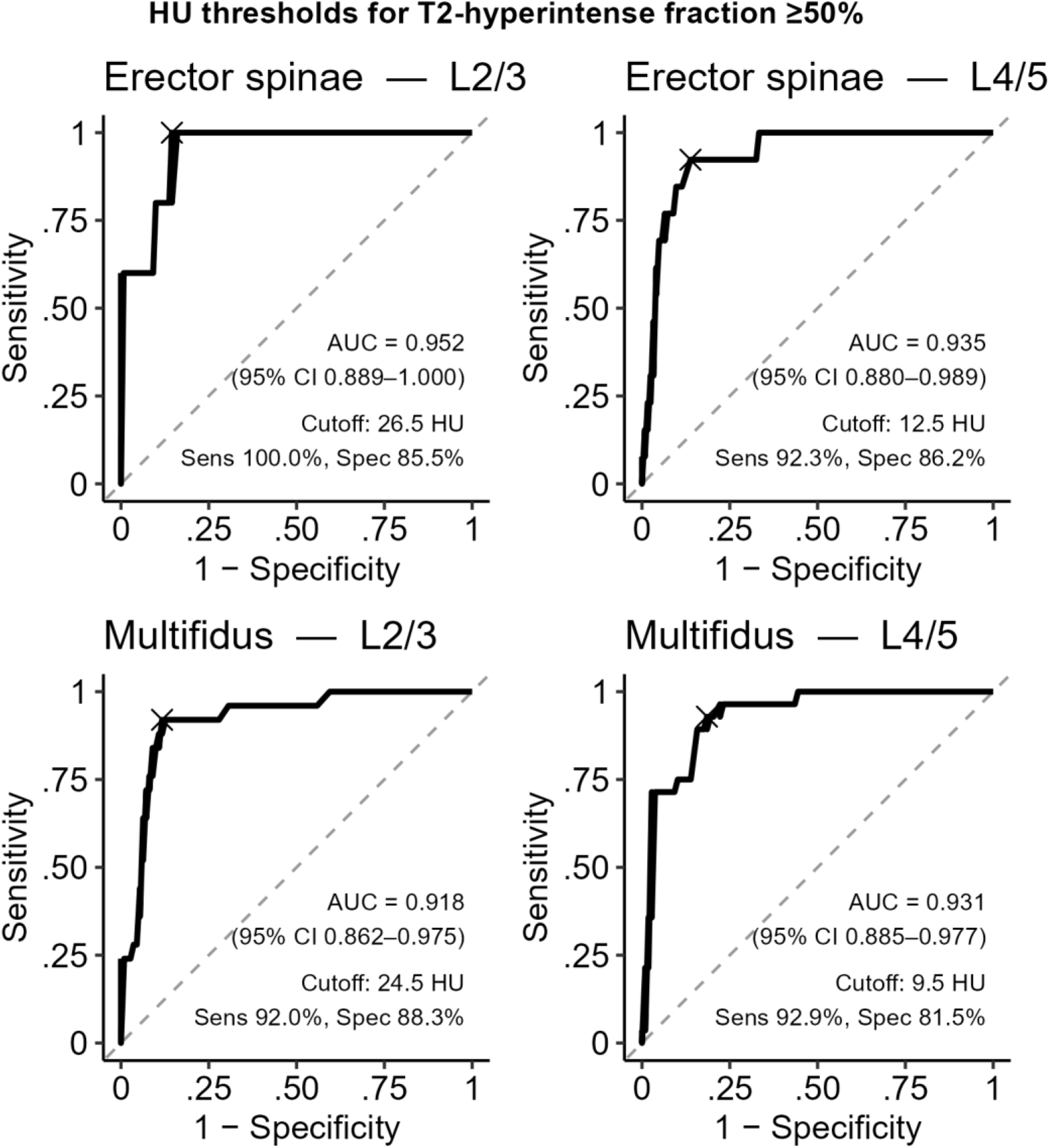
Level-specific receiver operating characteristic curves for detecting intramuscular T2-hyperintense area fraction ≥50%. Each analysis used the lower HU value side per patient, yielding n = 136 independent observations per panel. The “×” marks in each panel marks the Youden-optimal cutoff point. AUC with 95% confidence intervals (DeLong method), optimal HU cutoff, sensitivity, and specificity are annotated within each panel. HU, Hounsfield unit; AUC, area under the curve.

**Table 5.**
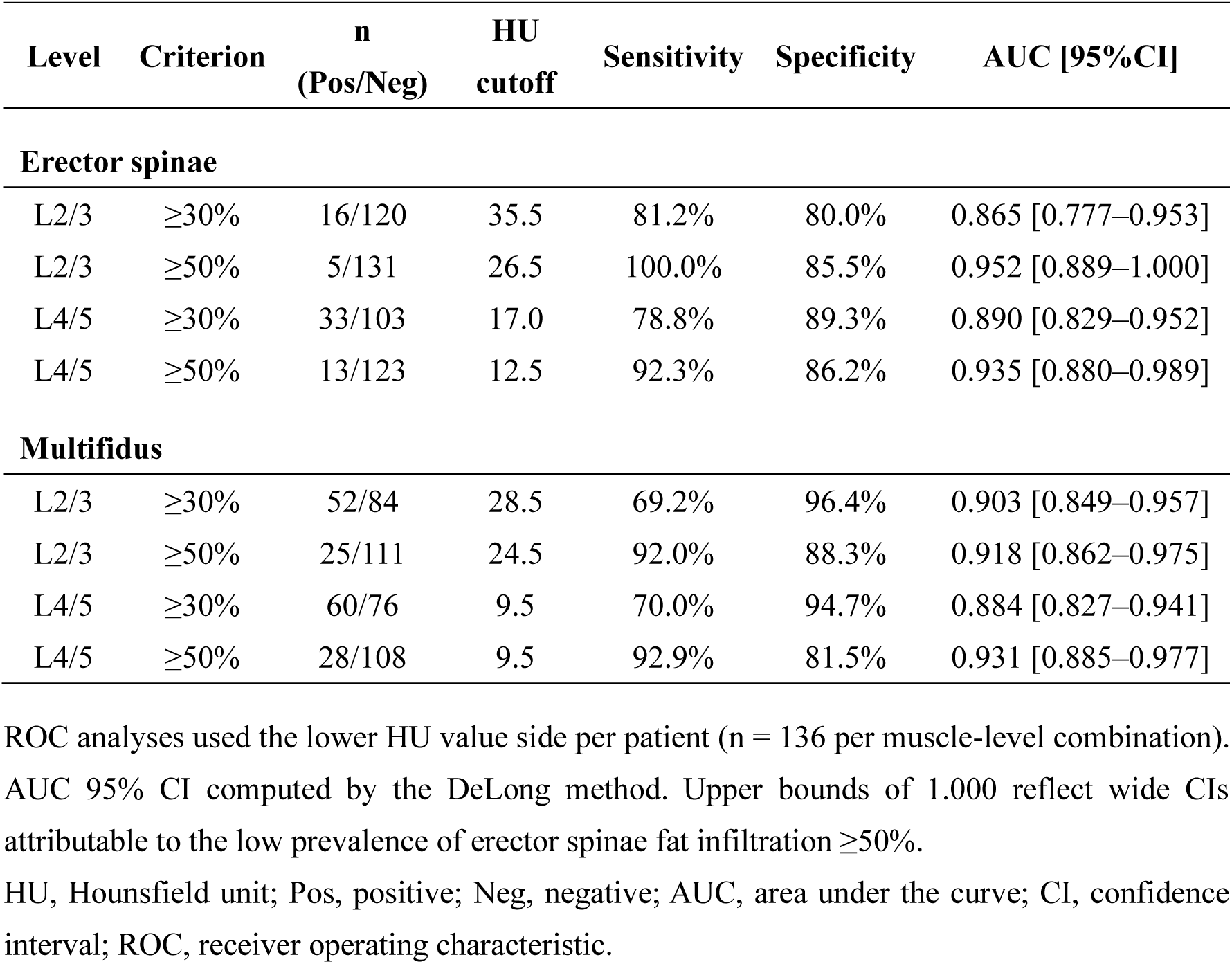
Level-specific HU thresholds for screening paraspinal muscle fat infiltration.

| Level | Criterion | n<br>(Pos/Neg) | HU<br>cutoff | Sensitivity | Specificity | AUC [95%CI] |
| --- | --- | --- | --- | --- | --- | --- |
| **Erector spinae** |  |  |  |  |  |  |
| L2/3 | ≥30% | 16/120 | 35.5 | 81.2% | 80.0% | 0.865 [0.777–0.953] |
| L2/3 | ≥50% | 5/131 | 26.5 | 100.0% | 85.5% | 0.952 [0.889–1.000] |
| L4/5 | ≥30% | 33/103 | 17.0 | 78.8% | 89.3% | 0.890 [0.829–0.952] |
| L4/5 | ≥50% | 13/123 | 12.5 | 92.3% | 86.2% | 0.935 [0.880–0.989] |
| **Multifidus** |  |  |  |  |  |  |
| L2/3 | ≥30% | 52/84 | 28.5 | 69.2% | 96.4% | 0.903 [0.849–0.957] |
| L2/3 | ≥50% | 25/111 | 24.5 | 92.0% | 88.3% | 0.918 [0.862–0.975] |
| L4/5 | ≥30% | 60/76 | 9.5 | 70.0% | 94.7% | 0.884 [0.827–0.941] |
| L4/5 | ≥50% | 28/108 | 9.5 | 92.9% | 81.5% | 0.931 [0.885–0.977] |

ROC analyses used the lower HU value side per patient (n = 136 per muscle-level combination). AUC 95% CI computed by the DeLong method. Upper bounds of 1.000 reflect wide CIs attributable to the low prevalence of erector spinae fat infiltration ≥50%.
HU, Hounsfield unit; Pos, positive; Neg, negative; AUC, area under the curve; CI, confidence interval; ROC, receiver operating characteristic.

## Discussion

In this retrospective study of 136 patients who underwent spine surgery with diverse diagnoses, we investigated whether paraspinal muscles could be assessed using HU measurements based on simple elliptical ROIs on routine preoperative CT scans. HU values measured using elliptical ROIs revealed a relatively good correlation with HU values measured using freehand ROIs. Furthermore, elliptical ROI-based HU values correlated with MRI-derived T2-hyperintense area fraction for both the erector spinae and multifidus muscles. A linear mixed-effects framework confirmed the robustness of this association after accounting for the hierarchical data structure.

The use of a simple elliptical ROI confers several practical advantages over that of freehand muscle segmentation. Manual freehand tracing of paraspinal muscles on CT is time-consuming and operator-dependent, limiting its wider clinical implementation.^18,19^ In this study, elliptical ROI placement required a mean of 4.0 ± 1.4 s per location, compared with 24.3 ± 13.2 s for freehand tracing, a six-fold reduction in measurement time. These findings were consistent with those of earlier studies demonstrating that simplified CT measurement methods reduce analysis time by up to 70% compared with full manual segmentation.^20^ Beyond speed, the elliptical approach eliminates the need for continuous cursor tracing along muscle boundaries, which is technically demanding and fatiguing during high-volume clinical workflows.^19^ In many PACS implementations, freehand ROI tools require uninterrupted single-stroke drawing without incremental adjustment, whereas elliptical ROIs can be repositioned and resized interactively until optimal placement is confirmed. Together with the good agreement between elliptical and freehand HU measurements (*R*, 0.776; ICC, 0.733), these characteristics make the elliptical approach substantially more suitable for routine clinical adoption.

The results of the current study have potentially critical implications for clinical practice. Given the diverse range of diagnoses in this study, diverse patterns of muscle degeneration were expected. ^21,22^ However, the HU–T2-fraction relationship was consistently strong across sex, age group, and pathological category. This diagnostic breadth supports the robustness of the CT HU approach. The fact that this simple measurement yielded comparable associations regardless of underlying diagnosis suggests that the observed relationship reflects genuine muscle physiology rather than reflecting disease-specific artifact. Therefore, the HU thresholds identified here may be applicable to a wide range of spinal surgery candidates. An elliptical ROI measurement requires only a few seconds of PACS work per muscle, and the reference values listed in Table 5 could serve as a simple, opportunistic screening method for patients who have already undergone routine CT scans.

Level-specific ROC analysis revealed that HU thresholds differ between the L2/3 and L4/5 levels. For the ≥30% fat infiltration criterion, the difference was approximately 20 HU for both erector spinae and multifidus. This discrepancy may be influenced by the prevalence of fat infiltration, which, reportedly, is higher in the lower lumbar spine,^23^ and shifts the Youden-optimal operating point. This finding cautions against applying a single level-independent HU threshold for the ≥30% criterion. For multifidus, the ≥50% criterion demonstrated smaller inter-level threshold differences and higher AUC than did the ≥30% criterion, making the ≥50% criterion a more informative screening target for this muscle. For erector spinae, however, the extremely low prevalence of ≥50% infiltration (positive n = 5–13 across analyses) rendered ROC-derived cutoffs statistically unreliable; nonetheless, these values should be considered hypothesis-generating only and must not be applied clinically without external validation.

Several conceptual considerations bear on the interpretation of these findings. The T2-hyperintense area fraction obtained via Otsu thresholding is an indirect surrogate for intramuscular fat infiltration; moreover, T2 hyperintensity may reflect edema, denervation atrophy, fibrous deposition, and other non-lipid pathological processes.^9,10^ Dixon-based fat-water separation provides a specific measurement of lipid protons^24^ and represents an optimal reference standard; however, Dixon sequences are not routinely available in standard lumbar MRI protocols and were not used in this study. Furthermore, thresholding of T2-weighted images cannot definitively distinguish intramuscular adipose tissue (between muscle fiber fascicles) from interfascicular and epimysial fat. In contrast, CT HU values measure X-ray attenuation averaged across the entire ellipse, integrating both the intramuscular and potentially perimuscular fat in proportion to their voxel contribution. Therefore, CT HU and MRI T2-hyperintense fraction measure related but non-identical constructs; the observed moderate-to-strong correlations reflect their conceptual equivalence, while acknowledging the distinction of this measurement.

The inter-observer ICC for elliptical ROI HU measurement was moderate (0.709), which was lower than that obtained for MRI T2-hyperintense fraction (0.857). This moderate agreement likely reflects the inherent challenge of determining the optimal ellipse placement on CT images, where muscle boundaries are less sharply defined than they are on MRI.^25^ Therefore, the reproducibility of elliptical ROI-based HU measurement highlights the need for standardized protocols, particularly because CT provides lower soft-tissue contrast than does MRI, increasing the risk of inadvertently including adjacent fascial or interfascicular structures within the ROI. When a muscle cross-section contains a focal region of distinctly different density, such as a fat pocket or fibrous band, ellipse placement materially affects the measured HU. We recommend that the ellipse be positioned to maximally inscribe the muscle cross-section, while actively avoiding peripheral focal abnormalities. A significant level effect was observed in both LME models, L4/5 HU was lower by 5.6 HU for the erector muscle and by 8.2 HU for the multifidus muscle, independently of T2-hyperintense fraction; this suggests that ROI contamination by interfascicular fat and fascial structures may be systematically greater at L4/5.

The selection of the L2/3 and L4/5 disc levels as the measurement sites reflects their direct clinical relevance in lumbar spinal surgery. The L4/5 disc space is the most commonly operated lumbar level, and the muscle quality at the operated level is directly relevant to local biomechanical conditions influencing screw fixation, fusion quality, and adjacent segment stress.^2^ The L2/3 level represents the upper lumbar region where the upper instrumented vertebra is frequently positioned in fusion constructs; degeneration at this level has been associated with proximal junctional kyphosis.^3^ Furthermore, measuring two levels allows distinction between focal and diffuse fat infiltration patterns, which may carry different prognostic implications. While the L3 vertebral body level has been widely adopted in oncology and sarcopenia research as a single representative cross-section,^18,26^ the disc-level approach used in this study aligns more naturally with the anatomical reference frame used in clinical spinal surgery planning.

This study had some limitations. The study sample comprised a single-center cohort of spinal surgery candidates; hence, generalizability to other institutions and non-surgical populations requires prospective validation. Although we made efforts to match the CT and MRI measurement levels, perfect anatomical correspondence cannot be guaranteed, particularly in patients with scoliosis or irregular disc morphology. CT and MRI were not always acquired on standardized scanners; variations in reconstruction parameters and MRI sequence design may have introduced measurement noise.^14^ The T2-hyperintense fraction is an imperfect surrogate for fat content as Dixon-based fat fraction is the preferred reference standard.^9^ Furthermore, this study did not directly evaluate the clinical outcomes; hence, the prognostic value of the identified HU thresholds should be confirmed prospectively. Notwithstanding these limitations, the finding that simple elliptical ROI measurements on clinical-grade CT correlate substantially with more labor-intensive MRI-based quantification—under realistic clinical conditions across a heterogeneous patient population—represents a meaningful step toward routine muscle quality screening in spinal surgery planning.

## Conclusion

The elliptical ROI-based HU measurement of lumbar paraspinal muscles demonstrated moderate-to-good inter-observer reliability and a strong association with MRI-derived intramuscular T2-hyperintense area fraction in both the erector spinae and multifidus muscles. HU thresholds differed substantially between the L2/3 and L4/5 spinal levels (∼20 HU for the ≥30% criterion), supporting the use of level-specific reference values. This approach integrates seamlessly into routine preoperative CT review using standard PACS software, without additional cost or radiation. The proposed thresholds may facilitate opportunistic screening of paraspinal muscle quality but require external validation before clinical implementation.

## Data Availability

All data produced in the present study are available upon reasonable request to the authors

## Acknowledgments

We would like to thank Editage (www.editage.com) for the English review.

## Ethical approval and informed consent

The study was conducted according to the guidelines of the Declaration of Helsinki and approved by the Institutional Review Board of Nagoya University School of Medicine (No. 2026-0034). The requirement for individual informed consent was waived because the study used only de-identified imaging data.

## Consent to participate

The requirement for individual informed consent was waived because the study used only de-identified imaging data.

## Consent for publication

Not applicable.

## Use of Generative AI

The authors disclose the use of Claude Sonnet 4.6 (Anthropic) during manuscript preparation. The tool was used to assist with manuscript preparation, including identifying potential methodological limitations and gaps in the analysis, and conducting literature searches. All outputs were carefully evaluated, verified, and edited by the authors. The authors are fully responsible for the accuracy and integrity of the work.

---

**Supplemental Figure 1.**
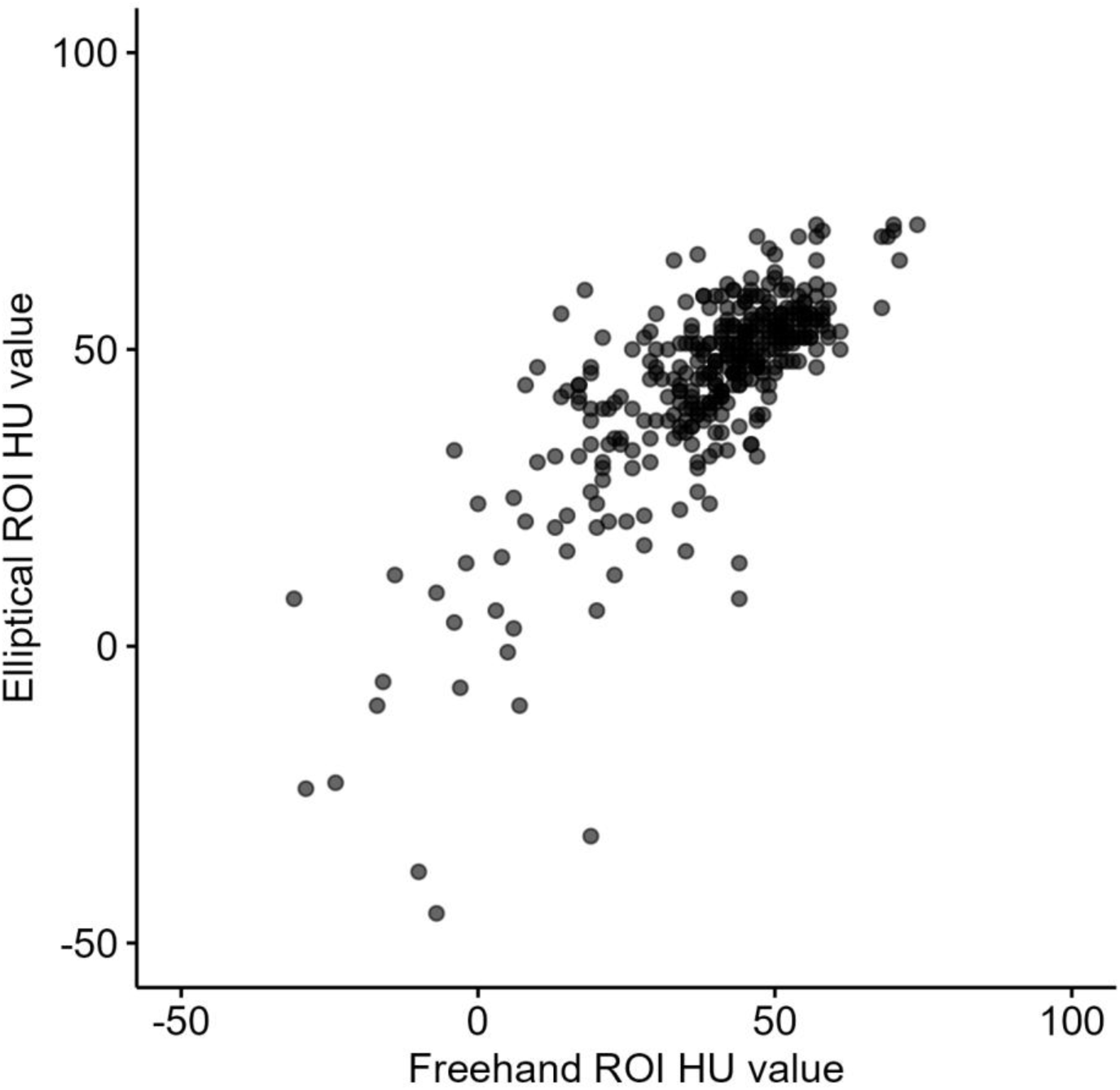
Correlation between HU values using freehand ROI and elliptical ROI. The correlation coefficient was 0.776 (95% confidence interval, 0.731–0.814). HU, Hounsfield unit; ROI, region of interest.

**Supplemental Figure 2.**
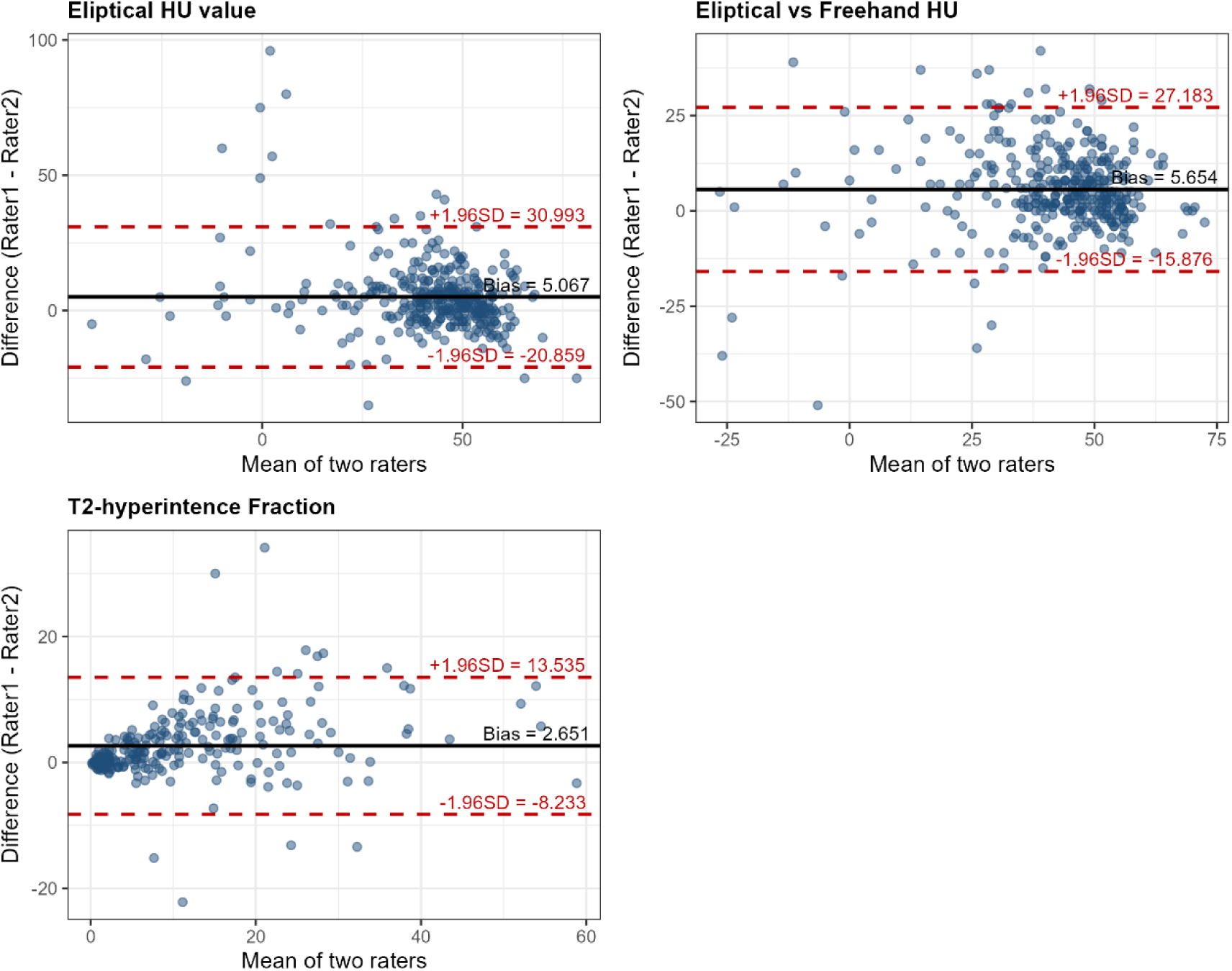
Bland-Altman plot of CT and MRI assessments. CT, computed tomography; MRI, magnetic resonance imaging; HU, Hounsfield unit.

